# Artificial intelligence-driven virtual tumor board enhances precision care in myelodysplastic syndromes

**DOI:** 10.64898/2026.03.26.26349088

**Authors:** David M Swoboda, Amy E DeZern, James T England, Sangeetha Venugopal, Thomas Kehoe, Brandon J Aubrey, Marco Gabriele Raddi, Angela Consagra, Jiasheng Wang, Jonathan Andreadakis, Gustavo Rivero, Maximilian Stahl, Amer M. Zeidan, Torsten Haferlach, Andrew M Brunner, Rena Buckstein, Valeria Santini, Matteo Giovanni Della Porta, Mikkael A Sekeres, Aziz Nazha

**Affiliations:** Tampa General Hospital Cancer Institute, Tampa FL; Sidney Kimmel Comprehensive Cancer Center, Johns Hopkins University, Baltimore MD; Sunnybrook Health Sciences Centre, Toronto Canada; University of Miami Sylvester Comprehensive Cancer Center, Miami FL; University of South Florida College of Medicine, Tampa FL; Massachusetts General Hospital, Boston MA; MDS Unit, DMSC-Hematology, University of Florence, AOU Careggi, Florence Italy; Department of Medical Biotechnologies, University of Siena, Siena, Italy; The Ohio State University Comprehensive Cancer Center, Columbus OH; Yale Comprehensive Cancer Center, Yale School of Medicine, Yale University, New Haven CT; MLL Munich Leukemia Laboratory, Munich Germany; Cancer Center, IRCCS Humanitas Research Hospital, Department of Biomedical Sciences, Humanitas University, Milan Italy; Sidney Kimmel Cancer Center, Thomas Jefferson University, Philadelphia PA

## Abstract

**Background:** Large language models (LLMs) perform well on standardized medical exam questions, but their reliability for complex hematology decision making is uncertain. We compared four general-purpose LLMs (GPT-4o, GPT-o3, Claude Sonnet 4, and DeepSeek-V3) with a Virtual MDS Panel (VMP), a coordinated multi-agent AI system in which domain-specialized, rule-bound software agents (WHO/ICC guidelines; IPSS-R/IPSS-M; NCCN) collaborate to generate tumor-board-level recommendations.

**Methods:** Each model generated diagnostic, prognostic, and treatment recommendations for 30 myelodysplastic syndrome cases. Nine international MDS experts from five institutions, blinded to model identity, completed 3,000 structured ratings using 5-point Likert scales for diagnosis, prognosis, and therapy and classified errors by severity.

**Results:** General-purpose LLMs achieved modest expert ratings (overall mean scores: 3.7 for GPT-o3, 3.2 for GPT-4o, 3.1 for DeepSeek, and 3.0 for Claude) and contained major factual errors in at least 24% of responses. The VMP increased the proportion of outputs rated 4 or higher to 87% (vs. 34-66% for general-purpose models), improved mean scores to 4.3 overall (4.3 for diagnosis, 4.4 for prognosis, and 4.1 for therapy), and reduced major errors to 8%.

**Conclusions:** In this blinded evaluation of 30 complex MDS cases, general-purpose LLMs produced clinically important errors at rates that raise safety concerns for autonomous hematology decision making. The VMP, a rule-bound, multi-agent architecture, approached expert-level accuracy supporting its potential role as an effective decision-support tool for MDS in the future.

## Introduction

Artificial intelligence (AI) has made substantial progress in healthcare over the past decade, driven by advances in deep learning, large-scale data availability, and increasing computational power.^1,2^ Early successes in medical imaging, pathology, and signal processing demonstrated that AI systems could match or exceed human-level performance in narrow, well-defined tasks.^3-6^ More recently, large language models (LLMs), such as ChatGPT, have expanded the scope of AI applications to complex cognitive domains, including clinical reasoning, medical documentation, and decision support.^7,8^

Notably, several contemporary LLMs have achieved passing or near-passing performance on standardized medical licensing and specialty board examinations, highlighting their ability to internalize vast amounts of biomedical knowledge and apply it in structured, exam-style settings.^9,10^

Parallel to these developments, the emergence of agentic AI, systems that can reason, understand context, and perform complex tasks with or without human supervision, has introduced a new paradigm for complex problem solving.^11^ Agentic architecture enables partitioning of tasks into discrete steps, iterative reasoning, and specialization across agents, mimicking aspects of human team-based decision-making.^12^ In healthcare, early applications of agentic AI have included multi-step clinical workflows, guideline navigation, literature synthesis, and preliminary diagnostic support.^13^ The utilization of this type of technology in this setting may be especially impactful for rural and underserved populations by extending multidisciplinary tumor board-level recommendations to patients who otherwise lack timely access to subspecialty expertise.^14,15^

Despite these advances, significant challenges remain in applying generative and agentic AI to real-world complex clinical scenarios.^16,17^ When presented with multiple-choice questions or clearly defined answer spaces, LLMs can leverage probabilistic pattern matching effectively, often yielding strong performance.^18,19^ However, real-world clinical problems, particularly in complex diseases, frequently lack a single correct answer, are influenced by incomplete or evolving evidence, and require complex clinical reasoning and judgment. In such settings, including oncology-related clinical scenarios, LLMs may generate superficially plausible but incomplete, inconsistent, or insufficiently justified recommendations, raising concern for medical error.^20-22^ This is increasingly relevant, as a recent study found that 21.5% of patients reported using ChatGPT for online health information, highlighting the importance of systematically evaluating these models and developing systems that are accurate and safe.^23^

To systematically evaluate this gap, we developed highly complex real-world clinical scenarios in myelodysplastic syndromes (MDS), a heterogeneous group of hematologic malignancies characterized by significant diagnostic complexity, rapidly evolving disease classification and prognostic systems, and individualized treatment strategies.^24-32^ We evaluated the performance of several leading foundation models, including GPT-4o, GPT-o3, DeepSeek-V3, and Claude Sonnet 4. We found that model responses often lacked disease-specific clinical reasoning, did not reconcile competing considerations, and omitted key nuances routinely recognized by experienced clinicians. Thus, we developed a highly scalable multi-agent virtual tumor board framework composed of specialized AI agents built to mimic multidisciplinary expert deliberation.

## Methods

### Virtual MDS Panel and Multiagent System Design

We designed a coordinated multi-agent AI system to emulate a multidisciplinary virtual tumor board for MDS (Figure 1). The system consists of domain-specialized agents designed to replicate expert clinical roles. The VMP comprises four collaborating agents:

**Figure 1:**
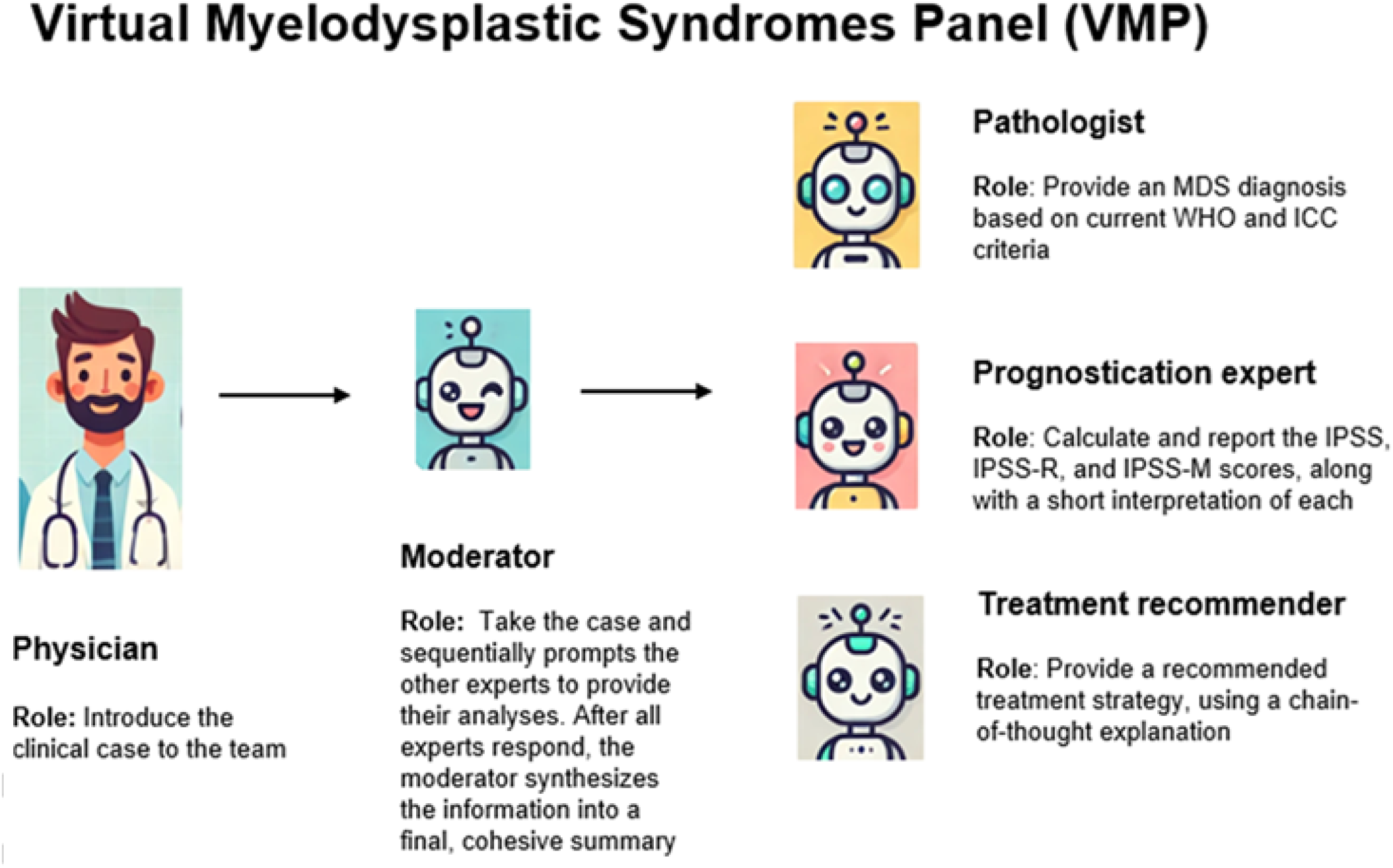
Architecture and Workflow of the Virtual Myelodysplastic Syndromes Panel (VMP). Schematic representation of the Virtual Myelodysplastic Syndromes Panel (VMP) workflow. A physician introduces a clinical case, which is coordinated by a moderator agent that sequentially prompts domain-specific expert agents. The pathology agent assigns an MDS diagnosis based on World Health Organization (WHO) and International Consensus Classification (ICC) criteria. The prognostication agent calculates IPSS, IPSS-R, and IPSS-M scores with brief interpretive summaries. The treatment recommendation agent proposes a management strategy. The moderator synthesizes all expert outputs into a final consolidated summary for clinical review. Source: Created by the authors.

1. **Moderator agent** receives clinician queries, routes tasks to the appropriate specialist agents, aggregates their outputs, and synthesizes a structured final response.
2. **Pathology agent** applies World Health Organization (WHO) and International Consensus Classification (ICC) criteria to case morphology, cytopenias, blasts, cytogenetics, mutations for diagnosis and classification.
3. **Prognostication agent** calculates and interprets IPSS, IPSS-R and IPSS-M risk categories and provides survival estimates.
4. **Therapy agent** generates evidence-based therapeutic recommendations based on established MDS clinical practice guidelines.

The agents operate in a coordinated, role-aware manner to enable collaborative reasoning analogous to a human tumor board. To reduce hallucinations (major factual errors), all agents were constrained to respond only when answers were explicitly supported by guideline-based evidence; otherwise, agents abstained from responding.

### Clinical Scenario Development and Model Benchmarking

We validated the minimum viable product (MVP), the most basic fully functional version of the VMP, by benchmarking model performance across complex, clinically realistic MDS scenarios. Thirty (n=30) high-fidelity synthetic case vignettes were developed by investigators to reflect real-world diagnostic and therapeutic complexity, integrating clinical presentation, hematologic parameters, morphology, blast percentage, cytogenetics (ISCN), and somatic gene variants required for WHO/ICC 2022 classification and IPSS-R/IPSS-M risk stratification. MDS were chosen as a prototypical highly complex oncologic disease requiring specialized clinical expertise, allowing robust model validation through expert consensus within a single, well-defined domain.

Cases were intentionally designed to test advanced clinical reasoning, including borderline blast thresholds, discordant cytopenias, IPSS-R lower-risk/IPSS-M higher-risk profiles, *NPM1*-mutant disease, and both frontline and relapsed treatment decisions. The reference standard (“ground truth”) for each case was defined by blinded consensus from an international expert panel.

For MVP evaluation, we benchmarked general-purpose large language models (GPT-4o, GPT-o3, Claude Sonnet 4, and DeepSeek-V3). Each model was evaluated using a single standardized prompt requesting diagnosis, prognosis and treatment recommendations. To prevent information leakage (carryover of case content or prior outputs across runs), each case was run in a new session and formatted using a fixed template with identical clinical inputs across models, with one output generated per model per case.

### Model evaluation and statistical analyses

Nine international MDS experts from five institutions, blinded to source and model identity, evaluated randomized, de-identified outputs, yielding 750 case-model evaluations and 3,000 structured ratings. Experts scored diagnosis, prognosis, and treatment on a 1–5 Likert scale (1=very inaccurate/incomplete; 5=completely accurate and detailed), with scores ≥”4 prespecified as acceptable, consistent with published frameworks.^33-35^ Overall performance was calculated as the unweighted average of the diagnosis, prognosis, and treatment scores. Factual accuracy was assessed separately and classified as no errors, minor inaccuracies (overall acceptable), or major factual errors or unsupported claims. Likert ratings (1–5) for diagnosis, prognosis, and treatment were averaged within institutions and summarized as mean ± SD, proportion of responses rated correct (score ≥”4) and major factual error rates. Pairwise comparisons between the Virtual MDS Panel (VMP) and each general-purpose model were performed using two-sided paired Wilcoxon signed-rank tests with Bonferroni correction, with effect sizes reported as rank-biserial correlation (RBC; r). Subgroup analyses compared the VMP with the pooled mean of the general LLMs using paired Wilcoxon tests, and between-group subgroup comparisons were evaluated using Mann–Whitney U tests. Intraclass correlation coefficients (ICCs) and 95% confidence intervals were calculated using a 2-way random-effects, mean-rating (k=5), consistency model to assess reliability of institution-averaged reviewer scores. Two-way ANOVA was used to estimate variance attributable to case, institution, and residual factors, and Pearson correlation coefficients were used to assess consistency across scoring domains. Analyses were performed in SAS version 9.4 and R version 4.5.2.

## Results

### Cohort Characteristics

We evaluated 30 synthetic vignettes (median age 69 years, range 45–85), of whom 17 (56.7%) were male. IPSS-R risk could be assigned to 28: very-low 2/28 (7.1%), low 11/28 (39.3%), intermediate 6/28 (21.4%), high 2/28 (7.1%), and very-high 7/28 (25.0%); thus, 19/28 (67.9 %) were lower-risk (very-low, low, intermediate) and 9/28 (32.1 %) higher-risk (high or very-high). Eighteen vignettes (60.0 %) were therapy-naïve and twelve (40.0%) therapy-exposed. Most were MDS (27/30, 90.0%); three were non-MDS (AML, CMML, CCUS). See Table 1 and Supplemental Table S1 for details.

**Table 1.**
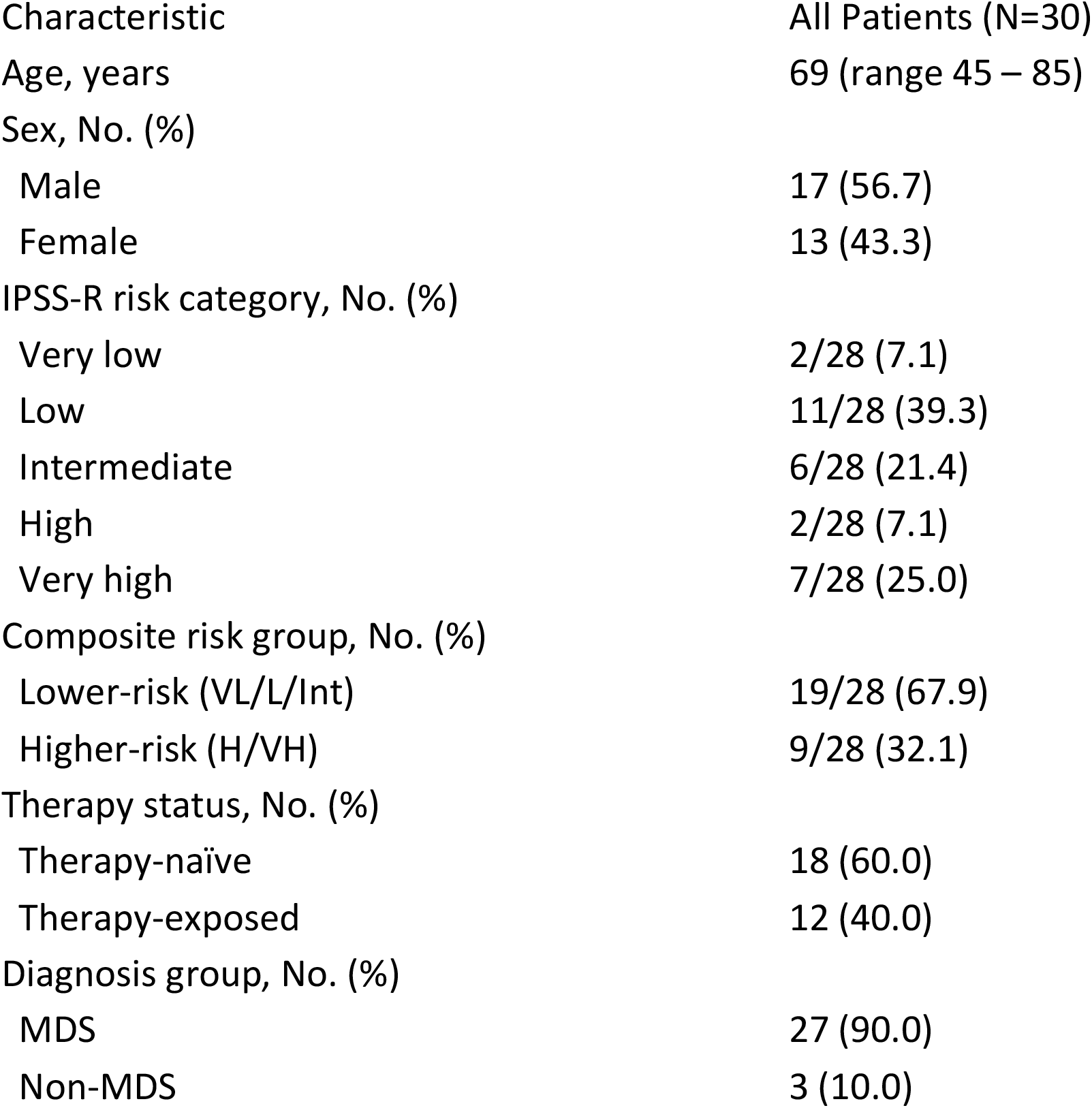
Baseline clinical characteristics of the evaluated case cohort. Shown are age, sex, IPSS-R risk category, composite risk group, therapy status, and diagnosis group for the 30 evaluated cases. IPSS-R risk was available for 28 cases. Lower-risk disease includes very low, low, and intermediate IPSS-R categories; higher-risk disease includes high and very high categories. Diagnosis groups were classified as myelodysplastic syndromes (MDS) versus non-MDS (AML, CMML, or CCUS). Detailed case-level characteristics are provided in Supplemental Table S1. Source: Created by the authors.

### Virtual MDS Panel

In blinded expert review, the general LLMs achieved acceptable performance in only 34-66% of model responses (66 % for GPT-o3, 41 % for GPT-4o, 38 % for DeepSeek, and 34 % for Claude).

Comparatively, the Virtual MDS Panel (VMP) was correct 87% of the time overall, including 88% correct diagnoses, 90% correct prognoses, and 83% correct therapy selections (Table 2). On the 1–5 Likert scale, the VMP’s mean scores were 4.3 for diagnosis, 4.4 for prognosis, and 4.1 for therapy (overall 4.3), each higher than GPT-o3 (3.7/3.7/3.6; overall 3.7), GPT-4o (3.1/3.2/3.3; overall 3.2), DeepSeek (3.0/3.1/3.2; overall 3.1), and Claude (2.8/3.0/3.2; overall 3.0); all differences were significant (Table 3). Performance distributions are shown in Figure 2. Major factual errors were lowest for the VMP (8%) and higher for GPT-o3 (24%), GPT-4o (25%), DeepSeek (31%), and Claude (32%) (Table 2).

**Table 2.** Comparative performance of evaluated models across diagnostic, prognostic, and treatment domains. Values represent the proportion of responses rated as correct (expert Likert score ≥”4) by domain and overall. Major factual errors (hallucinations) denote reviewer-flagged responses containing clinically significant inaccuracies. Source: Created by the authors.

| Model | Diagnosis | Prognosis | Treatment | Overall | Major Factual Error |
| --- | --- | --- | --- | --- | --- |
| VMP | 88.0 | 90.0 | 82.7 | 86.9 | 8.1 |
| GPT-o3 | 68.7 | 67.3 | 63.3 | 66.4 | 24.0 |
| GPT-4o | 38.0 | 41.3 | 43.3 | 40.9 | 24.8 |
| DeepSeek-V3 | 36.0 | 38.0 | 40.7 | 38.2 | 30.9 |
| Claude Sonnet 4 | 29.3 | 30.0 | 42.0 | 33.8 | 32.2 |

**Table 3.** Likert Performance Scores by Domain and Overall, Across Models. Values represent mean ± SD expert Likert scores (1–5) by domain and overall, summarized across institution-level case scores (one score per institution per case after within-institution averaging of reviewer ratings). †Two-sided paired Wilcoxon signed-rank tests (Bonferroni-adjusted) compare each model with the Virtual MDS Panel (VMP). Effect size reported as rank-biserial correlation (RBC; r). Values approaching 1 indicate that the Virtual MDS Panel consistently produced higher scores across paired case evaluations. Source: Created by the authors.

| Model | Diagnosis | Prognosis | Treatment | Overall | Overall p† | Effect size (RBC $r$)† |
| --- | --- | --- | --- | --- | --- | --- |
| VMP | 4.3 ± 0.9 | 4.4 ± 0.9 | 4.1 ± 1.0 | 4.3 ± 0.8 | — | — |
| GPT-o3 | 3.7 ± 1.1 | 3.7 ± 1.2 | 3.6 ± 1.2 | 3.7 ± 1.0 | <0.001 | 0.51 |
| GPT-4o | 3.1 ± 1.0 | 3.2 ± 1.0 | 3.3 ± 1.0 | 3.2 ± 0.8 | <0.001 | 1.0 |
| DeepSeek-V3 | 3.0 ± 1.1 | 3.1 ± 1.0 | 3.2 ± 1.0 | 3.1 ± 0.9 | <0.001 | 1.0 |
| Claude Sonnet 4 | 2.8 ± 1.2 | 3.0 ± 1.1 | 3.2 ± 1.1 | 3.0 ± 1.0 | <0.001 | 1.0 |

**Figure 2:**
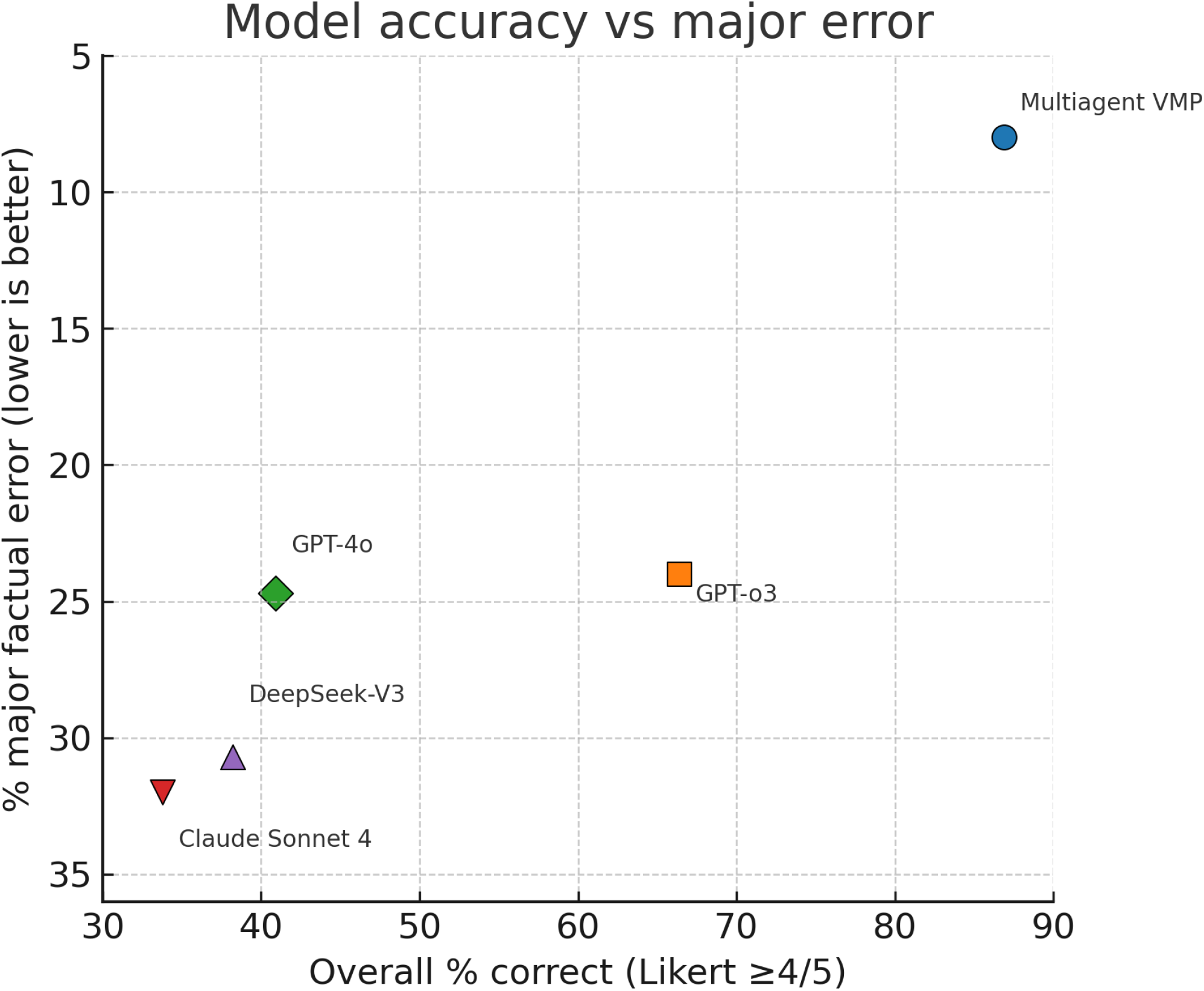
Comparative accuracy and major factual error rates across evaluated AI models. Scatter plot depicting overall accuracy (percentage of responses rated correct, Likert ≥”4) versus major factual error rate for each model. Points represent aggregate model performance across all cases and domains. Models positioned toward higher accuracy and lower error rates demonstrate more favorable overall performance. Source: Created by the authors.

### Subgroup analysis and case-level performance

The VMP performed similarly across cases involving lower-risk and higher-risk MDS, and among therapy-naïve and therapy-exposed patients, while consistently outperforming the single-model LLMs across these key clinical subgroups. Compared to the pooled average of GPT-o3, GPT-4o, DeepSeek, and Claude, the VMP achieved higher overall Likert scores by 0.9–1.4 points across IPSS-R risk groups, treatment-exposure strata, and MDS diagnoses (p<0.05 for all subgroups) (Table S2). Among-group comparisons for the VMP, such as age <70 vs ≥”70, male vs female, and lower risk (very low, low, intermediate) vs higher risk (high and very high), showed no statistically significant differences in performance (Table S3), indicating consistent scoring across demographic and clinical variables. For the single-model LLMs, performance was lower in lower-risk IPSS-R subgroups. At the case level, general LLMs scored low in cases involving AML and CMML, whereas the VMP maintained comparatively high scores across all case types (Table S4).

### Inter-rater reliability and score variance

Agreement across institutions, assessed using a two-way random-effects intraclass correlation coefficient (ICC[2,5]) on institution-level mean Likert scores, was moderate to good across all models (Diagnosis ICC=0.746; Prognosis ICC=0.782; Treatment ICC=0.694), consistent with the expected variability of expert assessment of complex, free-text clinical reasoning (Table S5).

Finally, we examined sources of variation in experts’ scores across model outputs. Most variance was attributable to case-level difficulty, reflecting scenarios with greater clinical ambiguity (e.g., incomplete or discordant data elements, competing management considerations, or limited consensus on optimal management) (Table S6). A smaller portion of variance reflected differences in scoring stringency across institutions and the remainder represented minor, unexplained variability. Despite inter-institutional differences, diagnostic, prognostic, and treatment scores were strongly correlated (r=0.58–0.72; all p<0.001), indicating that the scoring framework was internally consistent across domains (Table S7).

### Example case

A representative case is shown to illustrate how the Virtual MDS Panel (VMP) operates in practice, including sequential expert-agent outputs and moderator agent case synthesis and case summary. The example highlights the structured integration of classification (WHO/ICC guidelines), prognostic scoring (IPSS-R/IPSS-M), and therapy recommendations into a single, consolidated summary intended for clinician review. The full workflow trace and final synthesized output are shown in Figure 3.

## Discussion

Increasingly, patients are turning to widely available LLMs to assist them with understanding their own diagnosis, prognosis, and treatment.^23^ Health care providers, too, are incorporating AI, and specifically LLMs into their practices when evaluating patient cases.^36,37^ The accuracy of these approaches, especially across multiple unique LLMs, has not been well characterized for complex diseases such as MDS. Here, we showed that widely available general-purpose LLMs performed suboptimally on complex hematologic decision-making, reaching acceptable performance in only 34– 66% of responses. In contrast, the Virtual MDS Panel (VMP), a domain-focused, rule-bound multi-agent system, achieved near-expert performance (87% acceptable; mean score 4.3/5). Importantly in regard to safety, the VMP markedly reduced major factual or guideline-discordant errors, with only 8% of responses containing a major error compared with 24–32% among general-purpose LLMs. The ability to reduce errors is critical in medicine, where incorrect or inaccurate information can have dramatic implications on patients’ lives.

Importantly, VMP performance was consistent across demographic and clinical subgroups, with no statistically significant between-group differences including age, risk category, therapy exposure and non-MDS diagnoses. In contrast, general-purpose LLMs showed greater variability across case types and clinical contexts. Although the study was not powered to evaluate differences in subsets, these results suggest that a structured, domain-specific workflow may better preserve performance across different complex scenarios that require subspecialist interpretation.

Our findings also align with a growing body of literature highlighting important limitations of general-purpose LLMs in oncology. Prior studies across other cancer domains have shown that while such models often produce fluent and well-formed outputs, they frequently omit case-specific details or demonstrate reduced accuracy in advanced or molecularly complex disease.^20-22^ For example, in a breast cancer tumor board simulation, treatment recommendations generated by a general-purpose LLM were fully concordant with expert opinions in fewer than half of cases, and only approximately one-third of responses remained identical across repeat prompts.^20^ Large oncology benchmarks similarly demonstrate persistent clinically meaningful error rates even in advanced models^38^. These observations underscore the risks of deploying unmodified general-purpose LLMs in high-stakes, data-dense clinical environments such as hematology.

The superior performance of the VMP highlights the value of a constrained, knowledge-aware, multi-agent design.^13^ By breaking down clinical reasoning into specialized diagnostic, prognostic, and therapeutic tasks with explicit synthesis and cross-checking, the VMP mirrors the logic of a multidisciplinary tumor board. This helps mitigate several well-described failure modes of general-purpose LLMs, including inconsistent reasoning and inappropriate guideline application.^16,17^ In hematology, where management decisions depend on the accurate integration of morphology, cytogenetics, molecular features, and evolving guidelines, reducing serious factual or guideline-discordant errors is at least as important as maximizing overall performance scores.^1^

Despite the framework’s advanced reasoning and improved performance relative to general-purpose LLMs, several limitations warrant consideration. Although we evaluated highly complex synthetic cases designed to mimic real-world clinical practice, prospective evaluation on longitudinal patient records is needed to ensure the system can handle the full noise, documentation gaps, and operational constraints of routine clinical care. MDS served as an ideal test domain given its diagnostic and therapeutic complexity; however, validation across additional hematologic and solid tumor disease groups would strengthen generalizability. Additionally, we used a single standardized prompt to approximate typical real-world clinician use of general-purpose LLMs; however, performance for these models may improve with more tailored prompting and additional task-specific instructions. Finally, the LLM landscape is evolving rapidly: although we evaluated leading models available at the time of testing, model versions and capabilities change frequently, and newer models or updated releases may perform differently in these scenarios.

Nevertheless, these findings support the feasibility of AI systems that augment, rather than replace clinician judgment. The VMP is intended to function as a virtual tumor board: synthesizing complex information, cross-referencing established classification systems and prognostic frameworks, and presenting structured recommendations while leaving final decisions to the treating physician. Any real-world deployment must require rigorous safeguards for protected health information (PHI), including clear usage policies and built-in protections to prevent inadvertent disclosure (e.g., copy-pasted clinical notes). Such a human-in-the-loop paradigm is essential for maintaining safety, accountability, and clinician trust, particularly in a field where individual patient factors may defy algorithmic generalization.

In conclusion, the VMP outperformed general-purpose LLMs across diagnostic, prognostic, and therapeutic tasks while substantially reducing major factual and guideline-discordant errors. Taken together, these findings support domain-specific, knowledge-aware, multi-agent systems with clinician oversight as the most promising path forward for AI-enabled decision support in hematology and a likely foundation for future clinically deployable AI systems.

## Data Availability

Data Sharing Statement: Deidentified study data and analytic code will be shared upon reasonable request to the corresponding author, please contact

## Initial case information

A 72-year-old male with hypertension and type 2 diabetes has a WBC of 3.0 ×10^9^/L, ANC of 1.2 ×10^9^/L, hemoglobin of 7.2 g/dL,and platelets of 165 ×10^9^/L. Bone marrow evaluation demonstrated normocellularity at 40 % with >15 % ring sideroblasts and 2 % blasts. The morphology shows prominent erythroid dysplasia with abundant ring sideroblasts. Cytogenetic analysis showed a normal karyotype. Molecular profiling identified an SF3B1 mutation

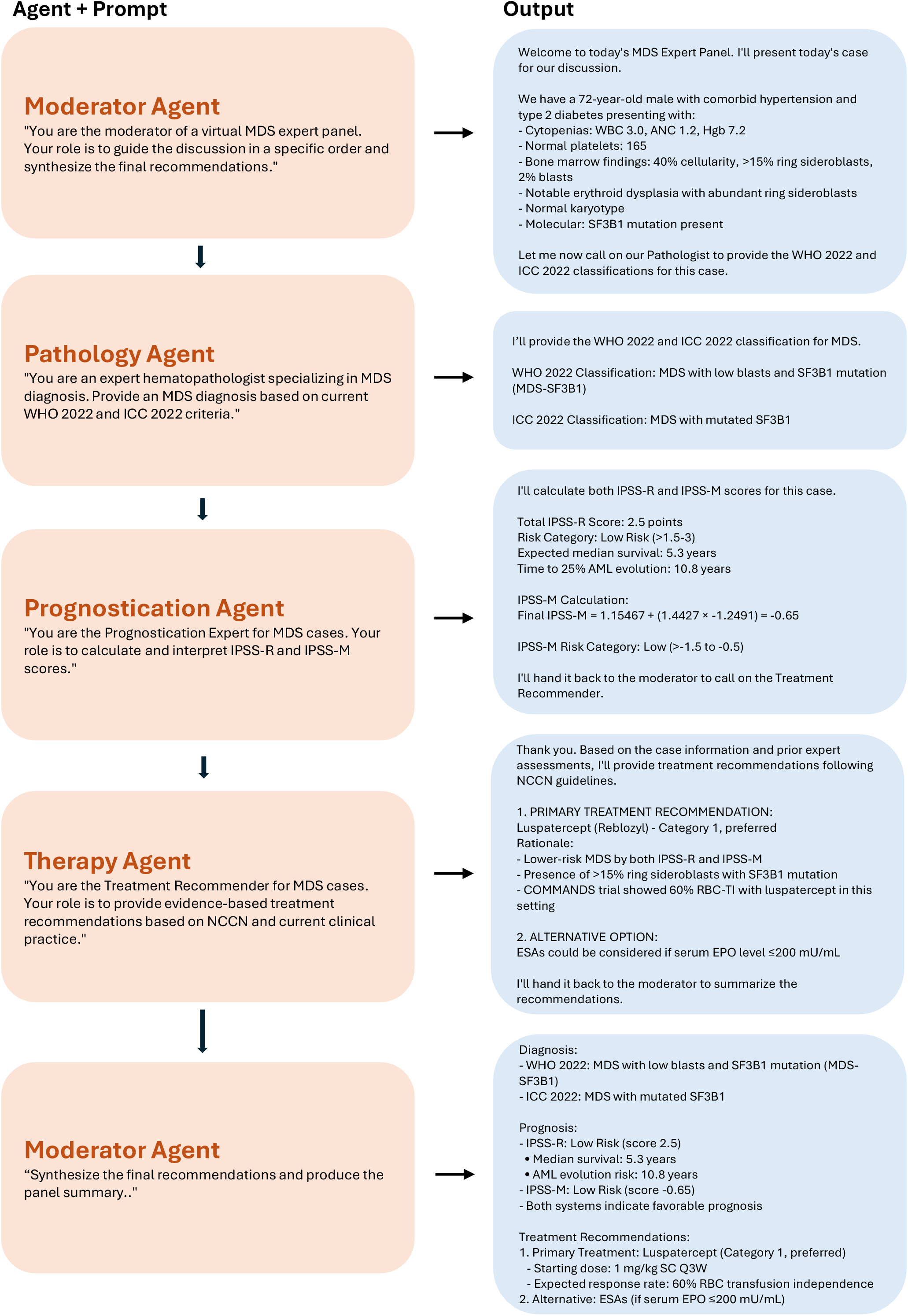

## Notes

### Competing Interest Statement

COI notes:
BMA: Receives royalty payments related to venetoclax from the Walter and Eliza Hall Institute of Medical Research (Melbourne, Australia).
RB: Research funding from Bristol Myers Squibb and Taiho; advisory board honoraria from Bristol Myers Squibb, Taiho, AbbVie, and Takeda; has served on trial steering committees for Takeda (formerly Keros) and Bristol Myers Squibb; and is a member of the Scientific Advisory Board of MDS-F.
AMB: Reports consulting fees from Novartis, AbbVie, Agios, Bristol Myers Squibb, Geron, i-Mab, Keros Therapeutics, Lava Therapeutics, Rigel, Sanofi, Syndax, Servier, and Takeda.
AC: No conflicts of interest to disclose.
AED: Has served as a consultant and/or in advisory roles for Bristol Myers Squibb, Novartis, Geron, Taiho, Keros, Agios, Takeda, UpToDate, CVS, and DynaMed.
JTE: Receives honoraria from Taiho, GSK, and Novartis.
TH: No conflicts of interest to disclose.
TK: No conflicts of interest to disclose.
AN: No conflicts of interest to disclose.
MGR: No conflicts of interest to disclose.
GR: No conflicts of interest to disclose.
VS: Has served on advisory boards for Ascentage, Bristol Myers Squibb, Geron, GSK, Jazz, Novartis, Servier, Pfizer, Alexion, Faron, and Takeda.
MAS: Has served on advisory boards for Bristol Myers Squibb, Rigel, Geron, and Agios.
MS: Served on advisory boards for Novartis, Kymera, Sierra Oncology, GSK, Rigel, Bristol Myers Squibb, Sobi, Syndax, Kura, and Servier; consulted for Boston Consulting Group, GLG, and The Dedham Group; participated in CME activities for Novartis, Curis Oncology, Haymarket Media, and Clinical Care Options; and is a member of the Medical Safety Monitoring Board for Takeda Pharmaceuticals.
MDP: No conflicts of interest to disclose.
DMS: Has served in consulting and/or advisory roles for Bristol Myers Squibb, Daiichi Sankyo, Geron, MorphoSys, and Syndax, and has participated in speakers bureaus for Bristol Myers Squibb, GSK, and Servier.
SV: No conflicts of interest to disclose.
JW: No conflicts of interest to disclose.
AMZ: Has participated in advisory boards, consulted, served on clinical trial committees, and/or received honoraria from AbbVie, Akesobio, Agios, Amgen, Astellas, BioCryst, Beigene, Boehringer Ingelheim, Celgene/Bristol Myers Squibb, Chiesi/Cornerstone Biopharma, Daiichi Sankyo, Dr. Reddy's, Epizyme, Faron, FibroGen, GSK, GlycoMimetics, Genentech, Gilead, Geron, Janssen, Jasper, Karyopharm, Kyowa Kirin, Keros, Kura, Novartis, Notable, Orum, Otsuka, Pfizer, Regeneron, Rigel, Seattle Genetics, Shattuck Labs, Schroedinger, Syros, Syndax, Servier, Takeda, Treadwell, Taiho, Vincerx, and Zentalis.

### Funding Statement

This study did not receive any funding.

