## Supplemental Materials for "Artificial intelligence-driven virtual tumor board enhances precision care in myelodysplastic syndromes"

**Abbreviations:** AML, acute myeloid leukemia; CCUS, clonal cytopenia of undetermined significance; CMML, chronic myelomonocytic leukemia; ICC, International Consensus Classification; IPSS-M, Molecular International Prognostic Scoring System; IPSS-R, Revised International Prognostic Scoring System; ISCN, International System for Human Cytogenomic Nomenclature; LLM, large language model; MDS, myelodysplastic syndromes; VMP, Virtual MDS Panel; WHO, World Health Organization.

[Table S5. Inter-Rater Reliability by Institution (ICC[2,5]) Across Domains.	14](#_Toc225152082)

### Supplemental Methods

#### Software and Implementation

The Virtual MDS Panel (VMP) was implemented in Python 3.13.7 within the Cursor development environment (v1.2.4). The orchestration layer used the Anthropic Python SDK to invoke a role-specialized, multi-agent workflow. Authentication was handled through an environment variable (ANTHROPIC_API_KEY) loaded at runtime.

All VMP agents (Moderator, Pathology, Prognostication Expert and Treatment Recommender) were powered by Anthropic Claude 3.5 Sonnet (model identifier: claude-3-5-sonnet-20241022). For each agent invocation, max tokens was set to 4000. Sampling parameters (e.g., temperature, nucleus sampling) were not explicitly set in the implementation and therefore used provider defaults.

#### Multi-agent Orchestration and Context Passing

VMP execution followed a fixed sequence: Moderator → Pathologist → Prognostication Expert → Treatment Recommender → Moderator (final synthesis). Each agent was prompted with a role-specific system message designed to emulate multidisciplinary tumor board deliberation. Agents were invoked via independent API calls, and prior agent outputs were passed forward as a plain-text transcript embedded in the user message (“Previous discussion”) to support iterative reasoning while maintaining role separation and a standardized ordering. The final Moderator synthesis produced a structured summary covering diagnosis (WHO 2022 and ICC 2022), prognosis (IPSS-R and IPSS-M), and treatment recommendations.

#### Hallucination Mitigation / Abstention Instructions

Agent system prompts included explicit instructions to avoid unsupported claims and to abstain when required inputs were missing or when recommendations were not guideline-supported. The orchestration layer did not implement external post-processing validation; hallucination mitigation was implemented through prompt-level constraints, role specialization, and structured synthesis.

#### Reliability and Error Handling

To improve robustness to transient API errors, the orchestration layer implemented exponential backoff retry logic (up to 5 attempts, with initial delay 2 seconds and doubling on subsequent retries) with structured logging of retry events and errors.

#### Comparator Model Access and Configuration

General-purpose comparator models (GPT-4o, GPT-o3, Claude Sonnet 4, and DeepSeek-V3) were benchmarked using their respective web user interfaces. For each case, all models received the same standardized prompt: “Please provide diagnosis, prognosis, treatment and clinical trial recommendations for the following case.” Each case–model pair was run in a new session to minimize carryover effects, and one output was collected per model per case. Repeated sampling was not performed; thus, within-model response variability was not assessed. Because web UI sessions do not consistently expose complete generation parameters, run dates and token/sampling settings were not programmatically captured. Accordingly, comparator outputs reflect platform-default configurations at the time of testing.

#### Web Access and Tools

For this MVP evaluation, the VMP was run without web browsing or external tool use; all outputs were generated using the models’ internal knowledge only.

#### Case inputs

Listed below are the actual 30 synthetic cases used in testing the 5 models.

1. A 72-year-old male with hypertension and type 2 diabetes has a WBC of 3.0 ×10⁹/L, ANC of 1.2 ×10⁹/L, hemoglobin of 7.2 g/dL, and platelets of 165 ×10⁹/L. Bone marrow evaluation demonstrated normocellularity at 40% with >15% ring sideroblasts and 2% blasts. The morphology shows prominent erythroid dysplasia with abundant ring sideroblasts. Cytogenetic analysis showed a normal karyotype. Molecular profiling identified an SF3B1 mutation.
2. A 58-year-old female with type 2 diabetes mellitus has a WBC of 3.5 ×10⁹/L, ANC of 1.8 ×10⁹/L, hemoglobin of 7.4 g/dL, and platelets of 120 ×10⁹/L. Bone marrow evaluation demonstrated slight hypocellularity at 25% with 1% blasts. The morphology shows marked erythroid dysplasia with megaloblastoid changes. Cytogenetic analysis revealed del(5q). Molecular profiling identified a TET2 mutation.
3. A 65-year-old male with a history of lymphoma treated with chemotherapy has a WBC of 2.0 ×10⁹/L, ANC of 0.8 ×10⁹/L, hemoglobin of 8.0 g/dL, and platelets of 45 ×10⁹/L. Bone marrow evaluation demonstrated hypercellularity at 75% with 7% blasts. The morphology shows multilineage dysplasia with dysgranulopoiesis and hypolobated megakaryocytes. Cytogenetic analysis showed monosomy 7. Molecular profiling identified biallelic TP53.
4. An 80-year-old female with coronary artery disease and chronic kidney disease is on luspatercept after ESA failure (EPO 530 mU/mL) and has a WBC of 3.8 ×10⁹/L, ANC of 1.8 ×10⁹/L, hemoglobin of 7.5 g/dL, and platelets of 150 ×10⁹/L. Bone marrow evaluation demonstrated 30% cellularity with 2% blasts. The morphology shows dyserythropoiesis with occasional dysplastic megakaryocytes. Cytogenetic analysis revealed del(20q). Molecular profiling identified a U2AF1 mutation.
5. A 54-year-old male with rheumatoid arthritis on methotrexate has a WBC of 9.0 ×10⁹/L, ANC of 6.0 ×10⁹/L, hemoglobin of 7.0 g/dL, and platelets of 200 ×10⁹/L; serum EPO 50 mU/mL. Bone marrow evaluation demonstrated 50 % cellularity with <5 % ring sideroblasts and 1% blasts. The morphology shows subtle erythroid dysplasia. Cytogenetic analysis revealed trisomy 8. Molecular profiling identified DNMT3A and TET2 mutations.
6. A 68-year-old female with a 40-pack-year smoking history has a WBC of 15.0 ×10⁹/L, ANC of 4.5 ×10⁹/L, hemoglobin of 9.8 g/dL, and platelets of 80 ×10⁹/L. Bone marrow evaluation demonstrated hypocellularity at 20 % with 11 % blasts. The morphology shows trilineage dysplasia with micromegakaryocytes, pseudo-Pelger–Huët neutrophils, and erythroid hypoplasia. Cytogenetic analysis showed a complex karyotype with del(5q), monosomy 7, and del(17p). Molecular profiling identified TP53 VAF 40% and RUNX1 mutations.
7. A 60-year-old male with obesity and nonalcoholic fatty liver disease has a WBC of 3.5 ×10⁹/L, ANC of 2.0 ×10⁹/L, hemoglobin of 7.2 g/dL, and platelets of 30 ×10⁹/L. Bone marrow evaluation demonstrated hypercellularity at 80 % with 9 % blasts. The morphology shows dysmegakaryopoiesis with small hypolobated megakaryocytes. Cytogenetic analysis revealed del(7q). Molecular profiling identified SRSF2 and IDH2 mutations.
8. A 70-year-old female with atrial fibrillation on anticoagulation has a WBC of 2.0 ×10⁹/L, ANC of 0.5 ×10⁹/L, hemoglobin of 8.5 g/dL, and platelets of 40 ×10⁹/L; serum EPO 200 mU/mL. Bone marrow evaluation demonstrated 35% cellularity with 18% blasts. The morphology shows pronounced multilineage dysplasia with significant dyserythropoiesis. Cytogenetic analysis showed del(5q). Molecular profiling identified TP53 VAF 25% and SF3B1 mutations.
9. A 62-year-old male with chronic obstructive pulmonary disease has a WBC of 4.5 ×10⁹/L, ANC of 3.0 ×10⁹/L, hemoglobin of 11.5 g/dL, and platelets of 140 ×10⁹/L. Bone marrow evaluation demonstrated 45% cellularity with 4% blasts. The morphology shows mild erythroid dysplasia with anisopoikilocytosis. Cytogenetic analysis showed a normal karyotype. Molecular profiling identified an SRSF2 mutation.
10. A 75-year-old female with diabetes and hypertension has a WBC of 5.0 ×10⁹/L, ANC of 2.5 ×10⁹/L, AMC 1.5×10⁹/L, hemoglobin of 7.8 g/dL, and platelets of 25 ×10⁹/L. Bone marrow evaluation demonstrated hypocellularity at 15% with 9% blasts. The morphology shows dysmegakaryopoiesis with giant platelet forms and erythroid hypoplasia. Cytogenetic analysis revealed del(5q) and del(20q). Molecular profiling identified an ASXL1, TET2, SRSF2 mutations.
11. A 45-year-old male with no significant comorbidities was treated with ESA (EPO 220 mU/mL) and has a WBC of 10.0 ×10⁹/L, ANC of 5.5 ×10⁹/L, hemoglobin of 7.5 g/dL, and platelets of 180 ×10⁹/L. Bone marrow evaluation demonstrated 60% cellularity with 2% blasts. The morphology shows subtle dyserythropoiesis and occasional dysplastic megakaryocytes. Cytogenetic analysis showed loss of the Y chromosome. Molecular profiling identified an SF3B1 mutation.
12. An 82-year-old female with Alzheimer’s disease, ECOG 4 has a WBC of 1.2 ×10⁹/L, ANC of 0.6 ×10⁹/L, hemoglobin of 9.0 g/dL, and platelets of 60 ×10⁹/L. Bone marrow evaluation demonstrated hypocellularity at 20% with 15% blasts. The morphology shows marked dysgranulopoiesis with pseudo-Pelger–Huët neutrophils. Cytogenetic analysis showed monosomy 7. Molecular profiling identified a TP53 mutation.
13. A 67-year-old male with gout has a WBC of 6.0 ×10⁹/L, ANC of 2.8 ×10⁹/L, AMC 0.2×10⁹/L hemoglobin of 7.8 g/dL, and platelets of 90 ×10⁹/L. Bone marrow evaluation demonstrated 55% cellularity with 3% blasts. The morphology shows mixed erythroid and megakaryocytic dysplasia. Cytogenetic analysis revealed trisomy 8 and del(5q). Molecular profiling identified ASXL1, DNMT3A, TET2 and U2AF1 mutations.
14. A 53-year-old female with no past medical history has a WBC of 1.0 ×10⁹/L, ANC of 0.4 ×10⁹/L, hemoglobin of 7.0 g/dL, and platelets of 150×10⁹/L; serum EPO 600 mU/mL. Bone marrow evaluation demonstrated 5% cellularity with 1% blasts. The morphology shows pronounced trilineage dysplasia with dysgranulopoiesis. Cytogenetic analysis showed a trisomy 8. Molecular profiling identified isolated TET2 mutation.
15. A 69-year-old male with metabolic syndrome has a WBC of 2.8 ×10⁹/L, ANC of 1.2 ×10⁹/L, hemoglobin of 9.5 g/dL, and platelets of 180 ×10⁹/L. Bone marrow evaluation demonstrated 65% cellularity with 1% blasts. The morphology shows absence of dysplasia. Cytogenetic analysis showed a normal karyotype. Molecular profiling identified a TET2 mutation.
16. A 77-year-old female with stage 3 chronic kidney disease and hypertension has a WBC of 3.2 ×10⁹/L, ANC of 0.9 ×10⁹/L, hemoglobin of 9.5 g/dL, and platelets of 50 ×10⁹/L. Bone marrow evaluation demonstrated hypocellularity at 15% with 4% blasts. The morphology shows multilineage dysplasia with micromegakaryocytes. Cytogenetic analysis revealed del(7q). Molecular profiling identified SRSF2 and RUNX1 mutations.
17. A 73-year-old male with no past medical history is post-luspatercept therapy (EPO 180 mU/mL) and has a WBC of 4.2 ×10⁹/L, ANC of 2.6 ×10⁹/L, hemoglobin of 8.7 g/dL, and platelets of 155 ×10⁹/L. Bone marrow evaluation demonstrated 70 % cellularity with 2% blasts. The morphology shows extensive erythroid and megakaryocytic dysplasia. Cytogenetic analysis showed trisomy 8. Molecular profiling identified ASXL1, TET2, U2AF1 mutations.
18. A 65-year-old female with nonalcoholic fatty liver disease has a WBC of 7.5 ×10⁹/L, ANC of 4.2 ×10⁹/L, hemoglobin of 7.2 g/dL, and platelets of 150 ×10⁹/L. EPO level 50. Bone marrow evaluation demonstrated 70% cellularity with 2% blasts. The morphology shows mild erythroid dysplasia. Cytogenetic analysis revealed del(20q). Molecular profiling identified SF3B1 and DNMT3A mutations. Patient has been on luspatercept for 1 year and now is becoming transfusion dependent.
19. A 85-year-old male with hypertension and hypothyroidism has a WBC of 4.1 ×10⁹/L, ANC of 2.5 ×10⁹/L, hemoglobin of 7.3 g/dL, and platelets of 190 ×10⁹/L; serum EPO 55 mU/mL. He was previously on ESA and then Luspatercept however is again transfusion dependent. Bone marrow evaluation demonstrated dyserythropoiesis, increased ringed sideroblasts >15%, no increased blast. Cytogenetic analysis was normal. Molecular profiling identified SF3B1 and IDH2 mutations.
20. A 65-year-old male with HIV CD4 350 has a WBC of 4.0 ×10⁹/L, ANC of 2.5 ×10⁹/L, AMC 0.2 ×10⁹/L, hemoglobin of 6.6 g/dL, and platelets of 208 ×10⁹/L. Erythropoietin 1566. Bone marrow evaluation demonstrated normocellular marrow at 40 % with no blasts. The morphology shows trilineage dyspoesis. Cytogenetic analysis showed normal karyotype. Molecular profiling identified IDH2, ASXL1 and SRSF2 mutations. He was previously given luspatercept and then enasidenib without improvement of anemia and still remains transfusion dependent.
21. A 63-year-old male with hypertension and mild osteoarthritis has a WBC of 2.2 ×10⁹/L, ANC of 0.6 ×10⁹/L, hemoglobin of 8.0 g/dL, and platelets of 20 ×10⁹/L. Bone marrow evaluation demonstrated 50% cellularity with 11% blasts. The morphology shows erythroid dysplasia without excess blasts. Cytogenetic analysis showed a normal karyotype. Molecular profiling identified an NPM1 mutation.
22. An 82-year-old male with chronic obstructive pulmonary disease and mild renal impairment has a WBC of 3.1 ×10⁹/L, ANC of 1.1 ×10⁹/L, hemoglobin of 8.2 g/dL, and platelets of 75 ×10⁹/L. He was previously treated with erythropoietin stimulating agent. Bone marrow evaluation demonstrated hypercellularity at 70% with 3% blasts. The morphology shows dysgranulopoiesis with pseudo-Pelger–Huët neutrophils. Cytogenetic analysis showed a normal karyotype. Molecular profiling identified an IDH1 mutation.
23. A 68-year-old female with type 2 diabetes and coronary artery disease has a WBC of 2.8 ×10⁹/L, ANC of 1.0 ×10⁹/L, hemoglobin of 7.5 g/dL, and platelets of 35 ×10⁹/L. She has received four cycles of decitabine with no hematologic improvement and bone marrow biopsy now shows 15% blasts (stable). The morphology shows persistent multilineage dysplasia with micromegakaryocytes. Cytogenetic analysis revealed del(7q). Molecular profiling identified ASXL1 and RUNX1 mutations.
24. A 72-year-old female with hypothyroidism has a WBC of 1.5 ×10⁹/L, ANC of 0.5 X 10⁹ AMC 0.1 ×10⁹/L, hemoglobin of 6.9 g/dL, and platelets of 20 ×10⁹/L. Bone marrow evaluation demonstrated hypercellularity at 90% with 9% blasts. The morphology shows marked erythroid and megakaryocytic dysplasia. Cytogenetic analysis showed a normal karyotype. Molecular profiling showing TET2, SRSF2 and FLT3 ITD. She has history of treatment with an ESA.
25. A 73-year-old male with no comorbidities has a WBC of 5.2 ×10⁹/L, ANC of 3.0 ×10⁹/L, hemoglobin of 9.0 g/dL, and platelets of 122 ×10⁹/L. EPO 20 mU/mL. Bone marrow evaluation demonstrated normocellular marrow involved by a myelodysplastic syndrome with predominantly megakaryocytic dysplasia, Blasts are not increased. Cytogenetic analysis showed a normal karyotype. Molecular profiling identified a CBL mutation.
26. A 79-year-old female with congestive heart failure and hypertension has a WBC of 1.0 ×10⁹/L, ANC of 0.2 ×10⁹/L, hemoglobin of 11.8 g/dL, and platelets of 160 ×10⁹/L. Bone marrow evaluation demonstrated 30% cellularity with 1% blasts. The morphology shows dysplastic neutrophils. Cytogenetic analysis revealed normal karyotype. Molecular profiling identified an ASXL1 mutation.
27. A 66-year-old male with prior pelvic radiation for prostate cancer has a WBC of 4.5 ×10⁹/L, ANC of 2.4 ×10⁹/L, hemoglobin of 9.9 g/dL, and platelets of 60 ×10⁹/L. Bone marrow evaluation demonstrated 45% cellularity with 12% blasts. The morphology shows multilineage dysplasia with dysgranulopoiesis. Cytogenetic analysis showed a monosomy 7. Molecular profiling identified RUNX1 mutations.
28. A 82-year-old male with hypertension and diabetes has a WBC of 3.8 ×10⁹/L, ANC of 2.0 ×10⁹/L, hemoglobin of 8.8 g/dL, and platelets of 56 ×10⁹/L. Bone marrow evaluation demonstrated normocellular marrow with 1% blasts. The morphology shows dysmegakaryopoiesis. Cytogenetic analysis revealed del(5q). Molecular profiling identified a TP53 VAF 19% and MSH6 mutations.
29. A 57-year-old male with alcohol use disorder has a WBC of 5.5 ×10⁹/L, ANC of 3.2 ×10⁹/L, hemoglobin of 10.7 g/dL, and platelets of 15 ×10⁹/L. Bone marrow evaluation demonstrated 60 % cellularity with 3 % blasts. The morphology shows megakaryocytic dysplasia. Cytogenetic analysis revealed del 20q. Molecular profiling identified a SRSF2 mutation.
30. An 85-year-old female with dementia and chronic kidney disease was exposed to ESA therapy (EPO 75 mU/mL) and has a WBC of 2.5 ×10⁹/L, ANC of 0.8 ×10⁹/L, hemoglobin of 7.1 g/dL, and platelets of 130 ×10⁹/L. Bone marrow evaluation demonstrated 15% cellularity with 2% blasts. The morphology shows pronounced dysgranulopoiesis with pseudo-Pelger–Huët cells. Cytogenetic analysis showed monosomy 7. Molecular profiling identified a TP53 mutation VAF 15%.

### Supplemental Tables

#### Table S1. Individual Case-Level Clinical and Molecular Characteristics.

For each case, age, sex, diagnosis, IPSS-R risk category, molecular alterations, and therapy status are shown. Source: Created by the authors from study data.

| Case | Age | Sex | Diagnosis | IPSS-R Risk | Molecular Profile | Therapy Status |
| --- | --- | --- | --- | --- | --- | --- |
| 1 | 72 | M | MDS | Low | SF3B1 | Therapy-naïve |
| 2 | 58 | F | MDS | Low | TET2 | Therapy-naïve |
| 3 | 65 | M | MDS | Very High | TP53 (biallelic) | Therapy-naïve |
| 4 | 80 | F | MDS | Low | U2AF1 | Therapy-exposed |
| 5 | 54 | M | MDS | Intermediate | DNMT3A, TET2 | Therapy-naïve |
| 6 | 68 | F | MDS | Very High | TP53 40%, RUNX1 | Therapy-naïve |
| 7 | 60 | M | MDS | Very High | SRSF2, IDH2 | Therapy-naïve |
| 8 | 70 | F | MDS | Very High | TP53 25%, SF3B1 | Therapy-naïve |
| 9 | 62 | M | MDS | Low | SRSF2 | Therapy-naïve |
| 10 | 75 | F | CMML | High | ASXL1, TET2, SRSF2 | Therapy-exposed |
| 11 | 45 | M | MDS | Very Low | SF3B1 | Therapy-exposed |
| 12 | 82 | F | MDS | Very High | TP53 | Therapy-naïve |
| 13 | 67 | M | MDS | Intermediate | ASXL1, DNMT3A, TET2, U2AF1 | Therapy-naïve |
| 14 | 53 | F | MDS | Intermediate | TET2 | Therapy-naïve |
| 15 | 69 | M | CCUS | N/A | TET2 | Therapy-naïve |
| 16 | 77 | F | MDS | Intermediate | SRSF2, RUNX1 | Therapy-naïve |
| 17 | 73 | M | MDS | Low | ASXL1, TET2, U2AF1 | Therapy-exposed |
| 18 | 65 | F | MDS | Low | SF3B1, DNMT3A | Therapy-exposed |
| 19 | 85 | M | MDS | Low | SF3B1, IDH2 | Therapy-exposed |
| 20 | 65 | M | MDS | Low | IDH2, ASXL1, SRSF2 | Therapy-exposed |
| 21 | 63 | M | AML | N/A | NPM1 | Therapy-naïve |
| 22 | 82 | M | MDS | Intermediate | IDH1 | Therapy-exposed |
| 23 | 68 | F | MDS | Very High | ASXL1, RUNX1 | Therapy-exposed |
| 24 | 72 | F | MDS | High | TET2, SRSF2, FLT3-ITD | Therapy-exposed |
| 25 | 73 | M | MDS | Low | CBL | Therapy-naïve |
| 26 | 79 | F | MDS | Very Low | ASXL1 | Therapy-naïve |
| 27 | 66 | M | MDS | Very High | RUNX1 | Therapy-exposed |
| 28 | 82 | M | MDS | Low | TP53 19%, MSH6 | Therapy-naïve |
| 29 | 57 | M | MDS | Low | SRSF2 | Therapy-naïve |
| 30 | 85 | F | MDS | Intermediate | TP53 15% | Therapy-exposed |

#### Table S2. Subgroup comparison of expert Likert scores for the Virtual MDS Panel (VMP) and pooled general-purpose LLMs.

Mean Likert scores (± SD), proportions of outputs rated ≥4, and mean score differences (ΔLikert) are shown by subgroup. ΔLikert denotes the difference in mean Likert score (VMP minus pooled LLM mean). The pooled LLM mean represents the average performance of GPT-o3, GPT-4o, DeepSeek-V3, and Claude Sonnet 4. p-values were calculated using paired Wilcoxon signed-rank tests. Source: Created by the authors from study data.

| Subgroup | Domain | VMP Mean ± SD | LLM Mean ± SD* | ΔLikert (VMP–LLM) | %≥4 VMP | %≥4 LLM* | p-value† |
| --- | --- | --- | --- | --- | --- | --- | --- |
| IPSS-R Lower-risk (n=19) | Overall | 4.2 ± 0.5 | 3.1 ± 0.4 | +1.02 | 74% | 0% | <0.001 |
| IPSS-R Higher-risk (n=9) | Overall | 4.5 ± 0.4 | 3.6 ± 0.4 | +0.87 | 89% | 22% | 0.004 |
| Therapy-naïve (n=18) | Overall | 4.3 ± 0.5 | 3.2 ± 0.6 | +1.03 | 83% | 11% | <0.001 |
| Therapy-exposed (n=12) | Overall | 4.3 ± 0.4 | 3.2 ± 0.3 | +1.03 | 75% | 0% | <0.001 |
| Diagnosis: MDS (n=27) | Overall | 4.3 ± 0.5 | 3.3 ± 0.4 | +0.99 | 81% | 7% | <0.001 |
| Diagnosis: non-MDS (n=3) | Overall | 4.2 ± 0.7 | 2.8 ± 0.7 | +1.43 | 67% | 0% | — |

**LLM mean = average of GPT-4o, GPT-o3, DeepSeek-V3, and Claude Sonnet 4. †Paired Wilcoxon p-values for case-level comparisons.*

#### Table S3. Between-group comparisons of overall Likert scores within demographic and clinical subgroups.

Mean overall Likert scores are shown for subgroup A versus subgroup B separately for the Virtual MDS Panel (VMP) and pooled general-purpose LLMs. p-values were calculated using Mann–Whitney U tests. Source: Created by the authors from study data.

| Subgroup | VMP | LLM |
| --- | --- | --- |
| Age <70 vs ≥70 | 4.20 vs 4.36 (p=0.550) | 3.28 vs 3.20 (p=0.208) |
| Male vs Female | 4.19 vs 4.39 (p=0.444) | 3.19 vs 3.32 (p=0.725) |
| Diagnosis MDS vs non-MDS | 4.28 vs 4.22 (p=1.000) | 3.30 vs 2.79 (p=0.167) |
| IPSS-R Lower vs Higher | 4.16 vs 4.45 (p=0.087) | **3.14 vs 3.58 (p=0.0195)** |
| Therapy Naïve vs Exposed | 4.27 vs 4.28 (p=0.644) | 3.24 vs 3.25 (p=0.706) |

#### Table S4. Case-Level Comparative Performance Scores Across Models.

Values represent mean Likert scores (1–5) assigned by expert reviewers for each model and case. The final column shows the mean score across all models for each case. Source: Created by the authors.

| Case | Claude Sonnet 4 | DeepSeek-V3 | GPT-4o | GPT-o3 | VMP | Overall (All Models Mean) |
| --- | --- | --- | --- | --- | --- | --- |
| 1 | 3.47 | 3.40 | 3.60 | 3.80 | 4.53 | 3.76 |
| 2 | 3.73 | 3.33 | 3.00 | 4.60 | 4.33 | 3.8 |
| 3 | 3.00 | 3.93 | 3.27 | 3.93 | 4.53 | 3.73 |
| 4 | 3.40 | 3.07 | 2.93 | 2.67 | 4.33 | 3.28 |
| 5 | 2.07 | 1.87 | 2.20 | 2.33 | 3.00 | 2.29 |
| 6 | 3.40 | 3.73 | 3.60 | 3.93 | 4.20 | 3.77 |
| 7 | 4.07 | 4.27 | 3.60 | 4.13 | 4.80 | 4.17 |
| 8 | 3.13 | 3.40 | 3.53 | 4.33 | 4.80 | 3.84 |
| 9 | 3.00 | 3.07 | 3.20 | 4.40 | 3.67 | 3.47 |
| 10 | 2.27 | 2.73 | 3.27 | 3.53 | 3.47 | 3.05 |
| 11 | 3.20 | 3.40 | 3.67 | 4.07 | 4.20 | 3.71 |
| 12 | 3.67 | 4.07 | 4.27 | 4.33 | 4.87 | 4.24 |
| 13 | 2.27 | 3.00 | 2.40 | 4.13 | 3.07 | 2.97 |
| 14 | 2.33 | 3.27 | 3.60 | 3.80 | 4.53 | 3.51 |
| 15 | 3.33 | 2.33 | 4.00 | 3.93 | 4.47 | 3.61 |
| 16 | 2.80 | 3.27 | 2.93 | 3.73 | 4.60 | 3.47 |
| 17 | 2.60 | 2.80 | 3.07 | 3.47 | 3.93 | 3.17 |
| 18 | 2.93 | 3.27 | 3.20 | 3.87 | 4.73 | 3.60 |
| 19 | 3.07 | 2.87 | 3.40 | 4.40 | 4.53 | 3.65 |
| 20 | 3.27 | 3.20 | 3.07 | 3.33 | 3.87 | 3.35 |
| 21 | 1.73 | 2.13 | 1.73 | 2.47 | 4.73 | 2.56 |
| 22 | 3.27 | 2.60 | 3.27 | 3.93 | 4.80 | 3.57 |
| 23 | 3.47 | 2.93 | 3.80 | 4.13 | 4.40 | 3.75 |
| 24 | 2.67 | 3.20 | 2.80 | 3.73 | 4.47 | 3.37 |
| 25 | 3.73 | 3.47 | 2.73 | 3.27 | 4.40 | 3.52 |
| 26 | 2.13 | 2.73 | 2.87 | 3.60 | 4.13 | 3.09 |
| 27 | 3.93 | 3.33 | 3.87 | 3.53 | 4.53 | 3.84 |
| 28 | 2.40 | 2.40 | 2.87 | 2.87 | 4.00 | 2.91 |
| 29 | 3.27 | 2.93 | 3.13 | 3.13 | 4.20 | 3.33 |
| 30 | 2.60 | 2.80 | 3.07 | 3.00 | 4.13 | 3.12 |

#### Table S5. Inter-Rater Reliability by Institution (ICC[2,5]) Across Domains.

ICC values were calculated from institutional mean Likert scores for diagnosis, prognosis, treatment, and across all domains combined using the Shrout–Fleiss two-way random-effects model. 95% confidence interval included in parentheses. Source: Created by the authors from study data.

| Model | Diagnosis | Prognosis | Treatment | Mean ICC Across Domains |
| --- | --- | --- | --- | --- |
| Claude Sonnet 4 | 0.239 (0.092, 0.435) | 0.254 (0.105, 0.451) | 0.377 (0.214, 0.569) | 0.290 |
| DeepSeek-V3 | 0.232 (0.087, 0.428) | 0.264 (0.113, 0.461) | 0.234 (0.088, 0.430) | 0.243 |
| GPT-4o | 0.318 (0.160, 0.514) | 0.356 (0.194, 0.550) | 0.151 (0.022, 0.340) | 0.275 |
| GPT-o3 | 0.338 (0.178, 0.533) | 0.167 (0.034, 0.358) | 0.127 (0.003, 0.311) | 0.211 |
| VMP | 0.210 (0.069, 0.405) | 0.127 (0.003, 0.312) | 0.207 (0.066, 0.402) | 0.181 |
| All Models Combined | 0.746 (0.567, 0.865) | 0.782 (0.629, 0.885) | 0.694 (0.479, 0.838) | 0.741 |

#### Table S6. Variance component analysis of expert Likert scores.

Two-way ANOVA was used to partition total variance into case-level, institutional, and residual components for each model and domain. Source: Created by the authors from study data.

| Model | Domain | Case Var % | Institution Var % | Residual Var % |
| --- | --- | --- | --- | --- |
| Claude Sonnet 4 | Diagnosis | 0.181 | 0.241 | 0.578 |
| Claude Sonnet 4 | Prognosis | 0.198 | 0.221 | 0.581 |
| Claude Sonnet 4 | Treatment | 0.305 | 0.192 | 0.503 |
| DeepSeek-V3 | Diagnosis | 0.188 | 0.189 | 0.623 |
| DeepSeek-V3 | Prognosis | 0.228 | 0.136 | 0.636 |
| DeepSeek-V3 | Treatment | 0.198 | 0.153 | 0.649 |
| GPT-4o | Diagnosis | 0.295 | 0.072 | 0.633 |
| GPT-4o | Prognosis | 0.312 | 0.124 | 0.564 |
| GPT-4o | Treatment | 0.140 | 0.076 | 0.784 |
| GPT-o3 | Diagnosis | 0.309 | 0.085 | 0.606 |
| GPT-o3 | Prognosis | 0.143 | 0.144 | 0.713 |
| GPT-o3 | Treatment | 0.098 | 0.229 | 0.673 |
| VMP | Diagnosis | 0.193 | 0.081 | 0.726 |
| VMP | Prognosis | 0.117 | 0.078 | 0.805 |
| VMP | Treatment | 0.190 | 0.083 | 0.727 |

#### Table S7. Correlation of expert Likert scores across scoring domains.

Pearson correlation coefficients (r) and corresponding p-values are shown for pairwise comparisons of diagnostic, prognostic, and treatment scores. Source: Created by the authors from study data.

| Domain 1 | Domain 2 | Pearson r | p-value |
| --- | --- | --- | --- |
| Diagnosis | Prognosis | 0.723 | < 0.0001 |
| Diagnosis | Treatment | 0.583 | < 0.0001 |
| Prognosis | Treatment | 0.660 | < 0.0001 |

#### Table S8. Reviewer Participation Summary.

The number of cases scored, models reviewed, and total ratings contributed by each reviewer are shown. For each model reviewed, four ratings were recorded: diagnosis, prognosis, treatment, and factual error assessment. Source: Created by the authors from study data.

| Reviewer | Institution | Cases Scored | Models Reviewed | Total Ratings |
| --- | --- | --- | --- | --- |
| AC | Florence | 6 | 5 | 120 |
| AED | Hopkins | 30 | 5 | 600 |
| AMB | MGH | 6 | 5 | 120 |
| BJA | MGH | 24 | 5 | 480 |
| JTE | Sunnybrook | 18 | 5 | 360 |
| MAS | Miami | 12 | 5 | 240 |
| RB | Sunnybrook | 12 | 5 | 240 |
| SV | Miami | 18 | 5 | 360 |
| VS | Florence | 24 | 5 | 480 |

### Supplemental Figures

#### Figure S1. Distribution of Likert Scores by Model.

Violin plots depict the distribution of expert ratings (Likert scale 1–5) for each model across all cases and domains. Central dashed lines indicate median scores, with quartiles shown where applicable. Higher scores reflect greater expert-rated accuracy and clinical usefulness. Source: Created by the authors from study data.
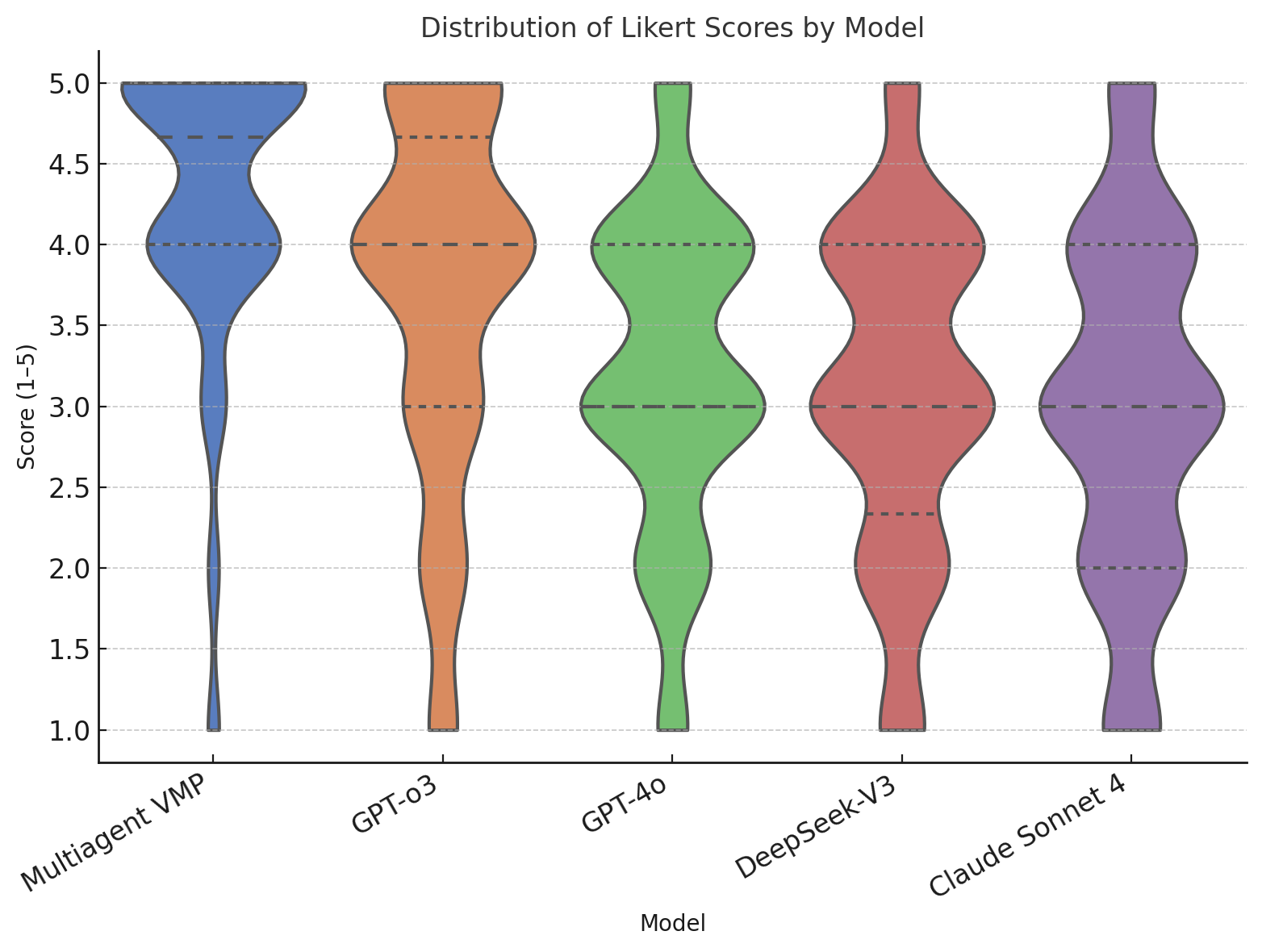

#### Figure S2. Domain-Specific Performance by Model (Diagnosis, Prognosis, Treatment).

Box-and-whisker plots show the distribution of Likert scores (1–5) for diagnostic, prognostic, and treatment recommendations generated by each model. Boxes represent interquartile ranges with median values indicated; whiskers denote the full range of observed scores. Scores ≥4 were prespecified as acceptable/correct. Source: Created by the authors from study data.

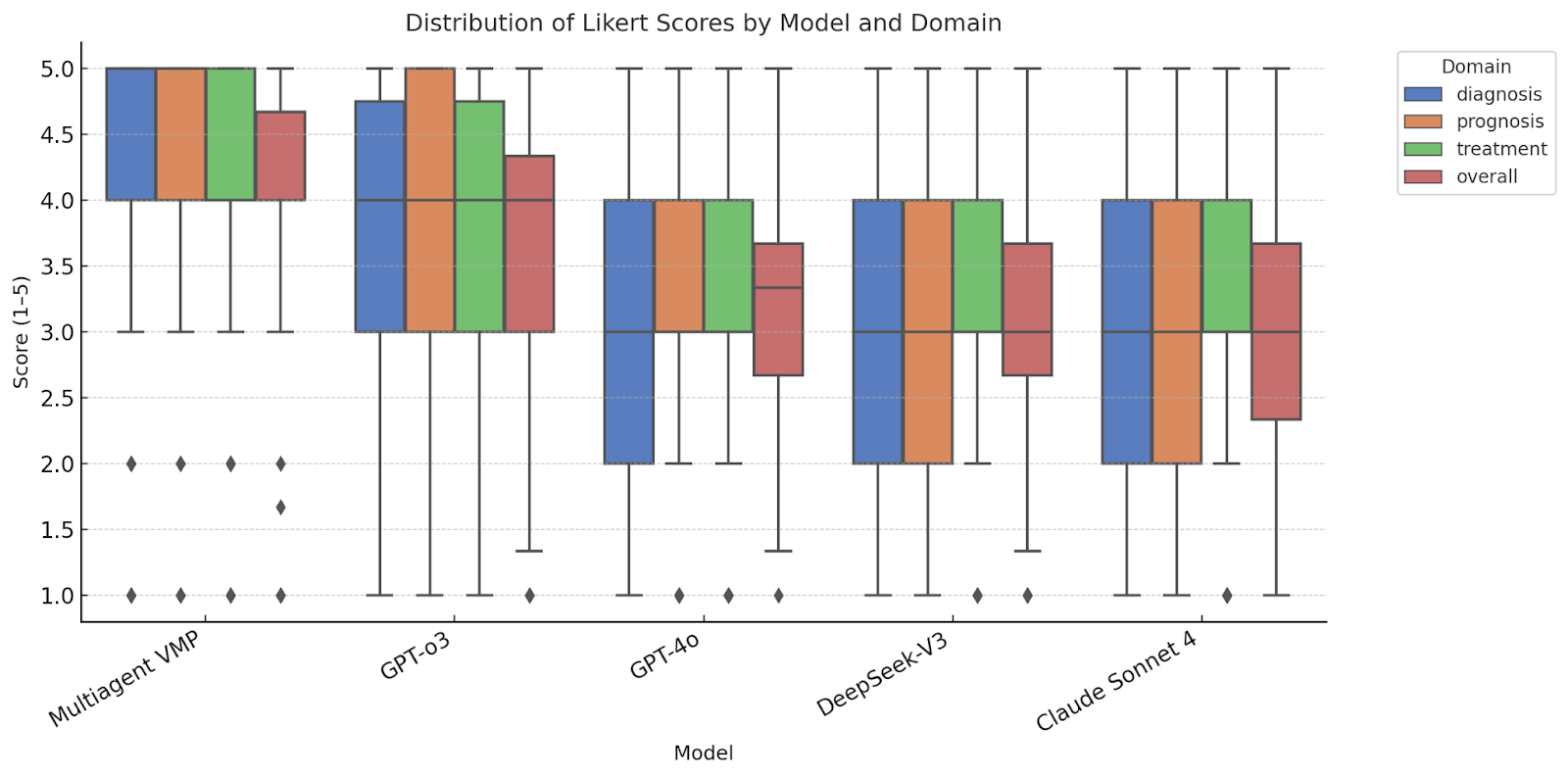

#### Figure S3. Consistent Performance Advantage of the Virtual MDS Panel Across Clinical Subgroups.

Forest plot showing differences in mean Likert scores (ΔLikert = Virtual MDS Panel minus pooled general-purpose LLM mean) across demographic and clinical subgroups. Positive values favor the Virtual MDS Panel. Points represent subgroup mean differences, and horizontal bars indicate 95% confidence intervals. Source: Created by the authors from study data.

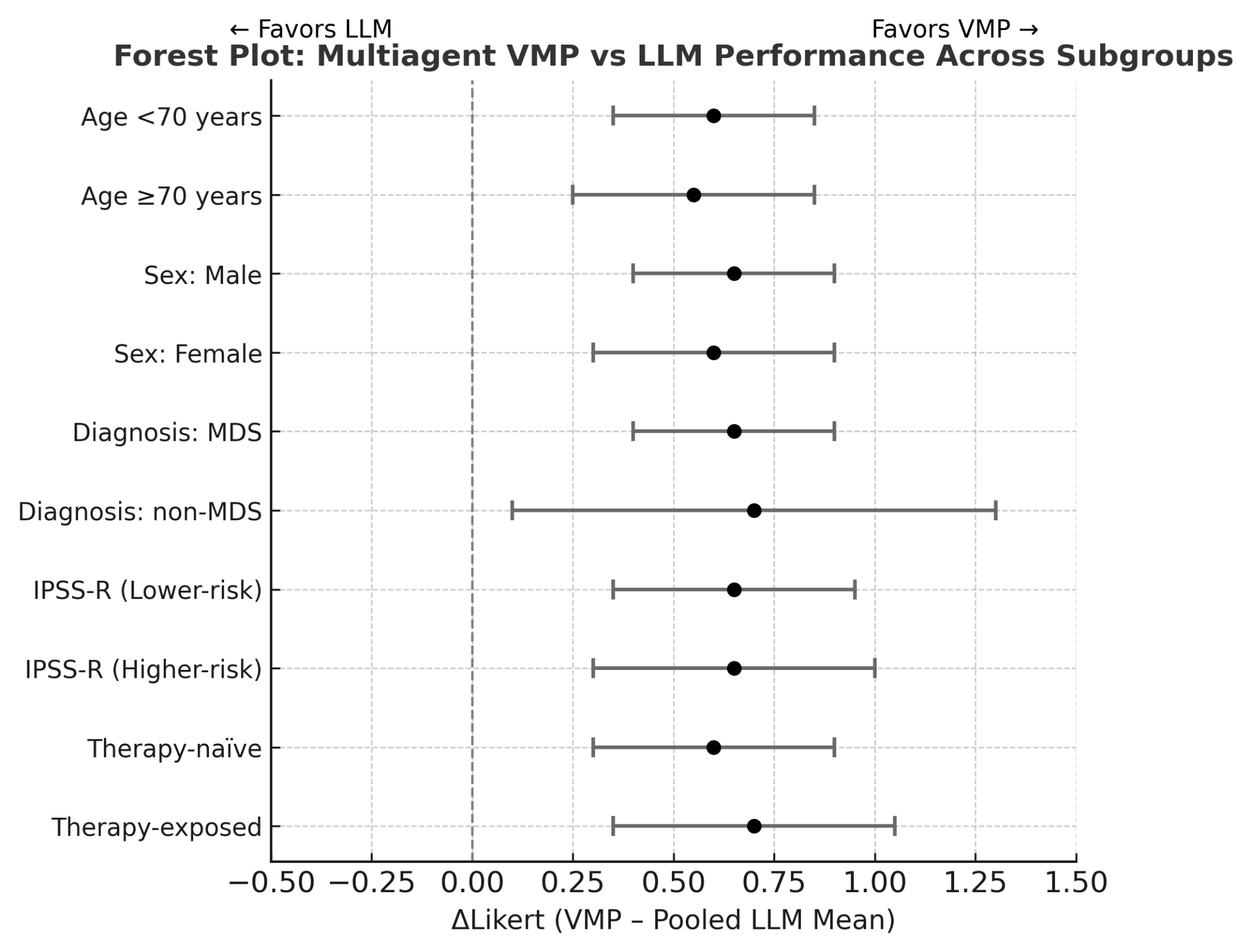
